# Participation in the nation-wide cervical cancer screening programme in Denmark during the COVID-19 pandemic: An observational study

**DOI:** 10.1101/2022.08.17.22278655

**Authors:** Tina Bech Olesen, Henry Jensen, Henrik Møller, Jens Winther Jensen, Marianne Waldstrøm, Berit Andersen

**Author notes:** **Corresponding author** Tina Bech Olesen, MSc, MSc, PhD, Project Manager, Resources & Innovation, The Danish Clinical Quality Program – National Clinical Registries (RKKP), Denmark.

## Abstract

**Background:** In contrast to most of the world, the cervical cancer screening programme continued in Denmark throughout the COVID-19 pandemic. We examined the cervical cancer screening participation during the pandemic in Denmark.

**Methods:** We included all women aged 23-64 years old invited to participate in cervical cancer screening from 2015-2021 as registered in the Cervical Cancer Screening Database combined with population-wide registries. Using a generalised linear model, we estimated prevalence ratios (PR) and 95% confidence intervals (CI) of cervical cancer screening participation within 90, 180 and 365 days since invitation during the pandemic in comparison with the previous years adjusting for age, year and month of invitation.

**Results:** Altogether, 2,220,000 invited women (in 1,466,353 individuals) were included in the study. Before the pandemic, 36% of invited women participated in screening within 90 days, 54% participated within 180 days and 65% participated within 365 days. At the start of the pandemic, participation in cervical cancer screening within 90 days was lower (pre-lockdown PR=0.58; 95% CI: 0.56-0.59 and 1^st^ lockdown PR=0.76; 95% CI: 0.75-0.77) compared with the previous years. A reduction in participation within 180 days was also seen during pre-lockdown (PR=0.89; 95% CI: 0.88-0.90) and 1^st^ lockdown (PR=0.92; 95% CI: 0.91-0.93). Allowing for 365 days to participation, only a slight reduction (3%) in participation was seen with slightly lower participation in some groups (immigrants, low education and low income).

**Conclusions:** The overall participation in cervical cancer screening was reduced during the early phase of the pandemic. However, the decline almost diminished with longer follow-up time.

**Funding:** The study was funded by the Danish Cancer Society Scientific Committee (grant number R321-A17417) and the Danish regions.

## INTRODUCTION

The COVID-19 pandemic is a global health crisis, which has caused extensive disruptions to the society and to the healthcare systems across the world. Population-wide restrictions (“lockdowns”) were imposed in most countries throughout the pandemic closing down schools and workplaces and restricting travel to reduce the transmission of COVID-19 and to limit the potential burden on the healthcare systems. Within the healthcare system, prioritisations and re-organisations were done to ensure sufficient capacity to take care of patients in need of hospitalisation due to COVID-19. The prioritisations within the healthcare system resulted in a temporary halting of the cervical cancer screening programme in most of the world. On the contrary, in Denmark the cervical cancer screening programmes remained open throughout the pandemic. At the same time; however, at the national televised press conferences, the health authorities asked the Danish population to stay at home if possible and concurrently, the Danish College of General Practitioners recommended general practitioners to postpone routine cervical smears during a four-week period in March/ April 2020 (1). Nevertheless, the cervical cancer screening programme continued – and invitations and reminders were sent out – throughout the pandemic in Denmark.

It is estimated that the disruptions to the cervical cancer screening programmes in high-income countries because of the pandemic could potentially increase cervical cancer cases by up to 5-6% and increase the number of cervical cancers detected at a higher stage (2). Disruptions to the cervical cancer screening programme may therefore be worrisome. Marked reductions in the number of women screened for cervical cancer during the early phase of the pandemic have been reported in many other countries (3-6), whilst the participation in cervical cancer screening during the pandemic in Denmark has not yet been described.

It is well known, that participation in cervical cancer screening is generally reduced among women of lower socio-economic status (7) and among immigrant women (8, 9). This divergence in participation may have been exacerbated during the COVID-19 pandemic. However, so far no studies have put spotlight on this.

In this large, population-based nationwide study, we examined the participation in cervical cancer screening during the COVID-19 pandemic in Denmark in comparison with the previous years. Moreover, we examined whether the participation in cervical cancer screening during the pandemic differed across population groups with different socioeconomic status.

## METHODS

### Setting

The study was set in Denmark, which has a population of approximately 5.8 million inhabitants (10). Denmark has a tax-funded healthcare system, with universal access to healthcare for all residents including national screening programmes for breast, cervical and colorectal cancer. The population-based administrative and health registries in Denmark can be linked through the unique personal identifier, assigned to all residents at birth or immigration (11, 12).

### The cervical cancer screening programme

In Denmark, all women aged 23-64 years old are invited to participate in cervical cancer screening every three years (women aged 23-49 years old) or every five years (women aged 50-64 years old) (13). The women receive an invitation letter (electronic letters via secure digital e-mail since 2018; however, women exempted from digital mail still receive ordinary mail) with an invitation to book an appointment with their general practitioner for a cervical screening test. Reminders to participate in cervical cancer screening are sent out to non-participants after 3 months and again after 6 months. The obtained samples are analysed for cytology and/or HPV at a pathology department. The outcome of the test is sent to the woman and her general practitioner.

### The COVID-19 pandemic in Denmark

In Denmark, three main waves of the COVID-19 pandemic have occurred that is, in the spring of 2020, in the winter of 2020/2021 and again in the winter of 2021/2022 (14).

In efforts to minimise the spread of the infection, population-wide restrictions (“lockdown”) were imposed in Denmark 11 March 2020 and subsequently, large parts of the society were closed down. Within the healthcare system, elective procedures were cancelled or postponed and resources were reallocated to take care of patients in need of hospitalisation because of COVID-19.

Extensive testing facilities were set up in Denmark from May 2020 providing COVID-19 tests free-of-charge to the whole population (15). Vaccination against COVID-19 began in December 2020 in Denmark and a high vaccination coverage has been achieved and by March 2022, approximately 81% of the population had received two doses and more than 61% had received three doses of the vaccine (16).

### Study population

The study population comprised all women aged 23-64 years old invited to participate in cervical cancer screening from 1 January 2015 to 30 September 2021, as registered in the Cervical Cancer Screening Database (17), which contain information on all women invited to participate in cervical cancer screening in Denmark since 2009. The Cervical Cancer Screening Database comprise population data from the Civil Registration System (11) including all persons with a permanent address in Denmark, cervical cancer cases are obtained from the Danish Cancer Register (18), cervical cytology samples are obtained from the Danish Pathology Register (19) and information on invitations and reminders is obtained from the invitation registration system.

We excluded invitations in women who died within 1 year since invitation (N=110), women who emigrated within 1 year since invitation (N=138), women residing in the Faroe Islands or Greenland (N=762), women with an unknown postal address (N=261), women who unregistered from the screening programme within 1 year since invitation (N=56,920) and invitations in women with missing information on region of residence (N=1,742) (Supplementary Figure 1).

### Exposure of interest

The exposure of interest was the COVID-pandemic in Denmark. The different phases of the pandemic were defined, in accordance with the governmental responses to the COVID-19 pandemic in Denmark, as follows:

- Pre-pandemic period: 1 January 2015 to 31 January 2020
- Pre-lockdown period: 1st February to 10 March 2020
- 1st lockdown: 11 March to 15 April 2020
- 1st re-opening: 16 April to 15 December 2020
- 2nd lockdown: 16 December 2020 to 27 February 2021
- 2nd re-opening: 28 February 2021 to 30 September 2021 (end of inclusion period)

We considered pre-lockdown and 1^st^ lockdown as the start of the pandemic.

### Outcome of interest

The main outcome of interest was participation in cervical cancer screening defined as having a cervical cancer screening test performed within 90, 180 and 365 days since invitation, respectively, among women invited to participate in the cervical screening programme.

### Explanatory variables

The following variables were examined independently: age, ethnicity, cohabitation status, educational level, disposable income and healthcare usage. Age was defined at the date of invitation, as registered in the Cervical Cancer Screening Database (17). From Statistics Denmark (10), we obtained information on ethnicity, marital status, educational level and level of income. Ethnicity was categorised as Danish descent, Western immigrant, Non-western immigrant and descendants of immigrants. Cohabitation status was categorised as single (i.e. living alone, divorced or not married), co-habiting/ co-living, and married (i.e. married or registered partnership) in accordance with Statistics Denmark (10). Education level was defined in accordance with the International Standard Classification of Education (ISCED) of the United Nations Education, Scientific and Cultural Organization (UNESCO) into short (ISCED level 1-2), medium (ISCED level 3-4) and long (ISCED level 5-8) (10). Income was defined as official disposable income depreciated to 2015 level and categorised into five quintiles. To indicate the level of healthcare use by each patient, we counted the total number of contacts to general practitioners, private practising medical specialists, physiotherapists, and chiropractors in the year for invitation as registered in the Danish National Health Service Register (20), which contain information on visits to primary healthcare (e.g., general practitioners and medical specialists) in Denmark since 1990. We categorised healthcare usage as rare (0-3 visits per year), low (4-6 visits per year), average (7-11 visits per year), high (12-18 visits per year) and frequent (≥19 visits per years).

### Statistical analyses

We examined characteristics of women invited to participate in cervical cancer screening during the study period. Thereafter, we examined the participation in cervical cancer screening within 90 days, 180 and 365 days since invitation among women invited to participate in screening per month and during the different phases of the pandemic overall and stratifying by the explanatory variables. Additionally, we examined time from invitation to participation in median number of days and interquartile interval (IQI) overall and during the pandemic phases.

Using a generalised linear model (GLM) with log link for the Poisson family with robust standard errors (SE), we estimated prevalence ratios (PR) and 95% confidence intervals (CI) of participation in cervical cancer screening within 90 days, 180 and 365 days, respectively, among women invited to participate in screening during the different phases of the pandemic overall and stratifying by the explanatory variables. Firstly, we calculated unadjusted analyses. Thereafter, the analyses were adjusted for month of invitation to allow for seasonality and year of invitation to take into account the underlying decreasing trend in participation in cervical cancer screening. Finally, the analyses were adjusted for age to take into account the effect of age on the other explanatory variables. All analyses were conducted using STATA version 17.0.

### Ethical considerations

The study is registered at the Central Denmark Region’s register of research projects (journal number 1-16-02-381-20). Patient consent is not required by Danish law for register-based studies.

## RESULTS

### Descriptive characteristics of the study population

Altogether, 2,220,000 invited women (in 1,466,353 individuals) were included in the study. The median age at invitation was 40 years (IQR=30-49 years), the majority of women (82.2%) were of Danish descent, 45.9% were married and 60.4% of women had a low educational level. The distribution of the descriptive characteristics was broadly similar throughout the study period (Table 1).

**Table 1.** Baseline characteristics of women invited to participate in cervical cancer screening in Denmark from 2015 to 2021.

|  | Pre-pandemic<br>(01Jan2015-31Jan2020) | Pre-lockdown<br>(01Feb2020-10Mar2020) | 1st lockdown<br>(11Mar2020-15Apr2020) | 1st re-opening<br>(16Apr2020-15Dec2020) | 2nd lockdown<br>(16Dec2020-27Feb2021) | 2nd re-opening<br>(28Feb2021-30Sep2021) | Total |
| --- | --- | --- | --- | --- | --- | --- | --- |
|  | N (%) | N (%) | N (%) | N (%) | N (%) | N (%) | N (%) |
| **Total** | 1641199 (73.9) | 41876 (1.9) | 31255 (1.4) | 223386 (10.1) | 69729 (3.1) | 212555 (9.6) | 2220000 (100.0) |
| **Age at invitation** |  |  |  |  |  |  |  |
| 23-29 years | 384272 (23.4) | 10223 (24.4) | 7731 (24.7) | 58569 (26.2) | 17624 (25.3) | 56816 (26.7) | 535235 (24.1) |
| 30-39 years | 412249 (25.1) | 10614 (25.3) | 7712 (24.7) | 56783 (25.4) | 17571 (25.2) | 54367 (25.6) | 559296 (25.2) |
| 40-49 years | 495153 (30.2) | 12690 (30.3) | 9233 (29.5) | 63872 (28.6) | 19051 (27.3) | 59366 (27.9) | 659365 (29.7) |
| 50-59 years | 246814 (15.0) | 6665 (15.9) | 5269 (16.9) | 34841 (15.6) | 11810 (16.9) | 31387 (14.8) | 336786 (15.2) |
| 60-64 years | 102711 (6.3) | 1684 (4.0) | 1310 (4.2) | 9321 (4.2) | 3673 (5.3) | 10619 (5.0) | 129318 (5.8) |
| Median (IQR) | 41 (31; 49) | 40 (30; 48) | 40 (30; 49) | 39 (30; 48) | 40 (30; 49) | 39 (30; 48) | 40 (30; 49) |
| **Ethnicity** |  |  |  |  |  |  |  |
| Danish descent | 1358106 (82.8) | 33975 (81.2) | 25778 (82.5) | 177501 (79.5) | 45340 (81.3) | 132376 (81.0) | 1773076 (82.2) |
| Descendant of immigrant | 31339 (1.9) | 918 (2.2) | 781 (2.5) | 5723 (2.6) | 1260 (2.3) | 3881 (2.4) | 43902 (2.0) |
| Western Immigrant | 87100 (5.3) | 2581 (6.2) | 1655 (5.3) | 15869 (7.1) | 3247 (5.8) | 9710 (5.9) | 120162 (5.6) |
| Non-western immigrant | 163638 (10.0) | 4375 (10.5) | 3027 (9.7) | 24176 (10.8) | 5928 (10.6) | 17535 (10.7) | 218679 (10.1) |
| **Cohabitation status** |  |  |  |  |  |  |  |
| Single | 529023 (32.3) | 13708 (32.8) | 10300 (33.0) | 76687 (34.4) | 3867 (35.1) | N/A | 633585 (32.5) |
| Cohabiting | 351991 (21.5) | 9225 (22.1) | 6829 (21.9) | 50379 (22.6) | 2369 (21.5) | N/A | 420793 (21.6) |
| Married | 759009 (46.3) | 18890 (45.2) | 14088 (45.1) | 96132 (43.1) | 4794 (43.5) | N/A | 892913 (45.9) |
| **Educational level (ISCED)** |  |  |  |  |  |  |  |
| ISCED15 level 1-2 | 960324 (60.6) | 24481 (59.3) | 18667 (60.6) | 129791 (59.3) | 40565 (60.1) | 122230 (60.4) | 1296058 (60.4) |
| ISCED15 level 3-4 | 393390 (24.8) | 10185 (24.7) | 7539 (24.5) | 52354 (23.9) | 16772 (24.8) | 48870 (24.1) | 529110 (24.7) |
| ISCED15 level 5-8 | 231157 (14.6) | 6589 (16.0) | 4600 (14.9) | 36716 (16.8) | 10173 (15.1) | 31395 (15.5) | 320630 (14.9) |
| **Disposable income** |  |  |  |  |  |  |  |
| Lowest quintile | 322307 (19.9) | 7419 (18.2) | 5869 (19.0) | 43920 (20.4) | 12715 (18.6) | 39486 (19.3) | 431716 (19.8) |
| Second quintile | 334113 (20.6) | 7611 (18.7) | 5878 (19.0) | 40851 (19.0) | 12137 (17.8) | 36480 (17.9) | 437070 (20.0) |
| Third quintile | 338563 (20.9) | 8066 (19.8) | 5850 (18.9) | 40863 (19.0) | 11738 (17.2) | 34157 (16.7) | 439237 (20.1) |
| Fourth quintile | 326174 (20.1) | 8604 (21.1) | 6449 (20.9) | 43279 (20.1) | 14025 (20.5) | 40367 (19.8) | 438898 (20.1) |
| Highest quintile | 301566 (18.6) | 9067 (22.2) | 6878 (22.2) | 46484 (21.6) | 17742 (26.0) | 53702 (26.3) | 435439 (20.0) |
| **Healthcare usage** |  |  |  |  |  |  |  |
| Rare | 319960 (19.5) | 8519 (20.3) | 5985 (19.1) | 47619 (21.3) | 13413 (19.2) | 43973 (20.7) | 439469 (19.8) |
| Low | 366807 (22.3) | 9020 (21.5) | 6774 (21.7) | 48280 (21.6) | 15181 (21.8) | 45102 (21.2) | 491164 (22.1) |
| Average | 347589 (21.2) | 8758 (20.9) | 6637 (21.2) | 46608 (20.9) | 14364 (20.6) | 44024 (20.7) | 467980 (21.1) |
| High | 299358 (18.2) | 7583 (18.1) | 5727 (18.3) | 40091 (17.9) | 12967 (18.6) | 38760 (18.2) | 404486 (18.2) |
| Frequent | 307485 (18.7) | 7996 (19.1) | 6132 (19.6) | 40788 (18.3) | 13804 (19.8) | 40696 (19.1) | 416901 (18.8) |
| **Time from invitation to participation, median (IQI)** | 94 (42; 200) | 120 (72; 207) | 122 (53; 201) | 86 (36; 161) | 69 (29; 133) | 51 (28; 101) | 89 (39; 184) |

IQI= interquartile interval; ISCED=International Standard Classification of Education

### Participation during the COVID-19 pandemic

Figure 1 shows the participation in cervical cancer screening within 90, 180 and 365 days throughout the study period. Before the pandemic, approximately 36% of women participated in cervical cancer screening within 90 days, 54% of women participated within 180 days and 65% of women participated within 365 days (Supplementary Tables 1-3).

**Figure 1.**
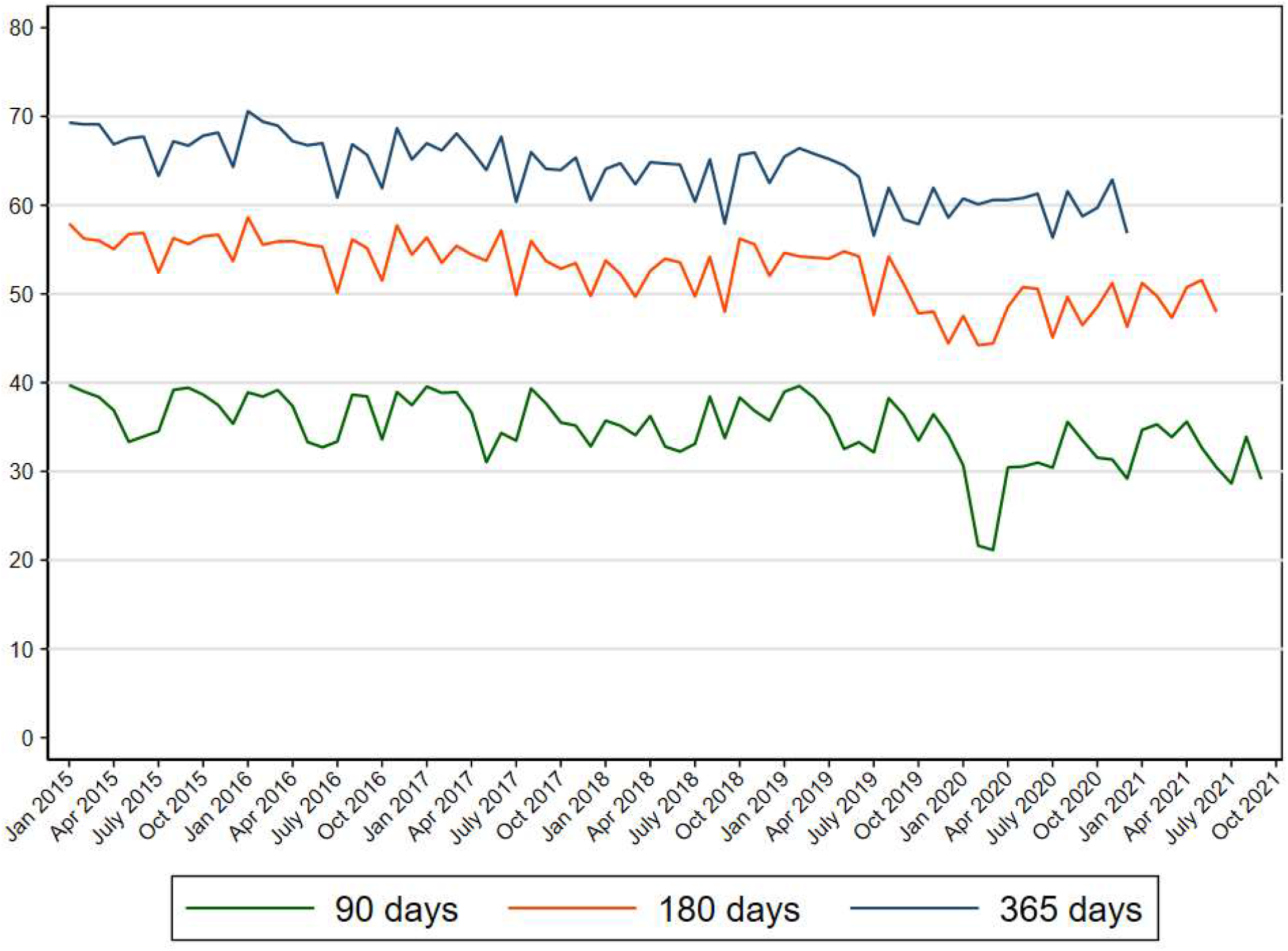
Participation in cervical cancer screening in Denmark within 90, 180 and 365 days since invitation from 2015 to 2021.

In March and April 2020, the participation in cervical cancer screening within 90 days dropped markedly to approximately 20% after which the participation resumed to normal levels (Figure 1). This was also reflected in a prevalence ratio (PR) of 0.58 (95% CI: 0.56-0.59) during pre-lockdown and a PR of 0.76 (95% CI: 0.75-0.77) during 1^st^ lockdown resuming to PRs of 0.96-0.99 throughout the rest of the study period (Supplementary Table 4).

A reduction in the participation in cervical cancer screening within 180 days was also observed among women invited at the start the pandemic (Figure 1) reflected in a PR of 0.89 (95% CI: 0.88-0.90) during pre-lockdown and a PR of 0.92 (95% CI: 0.91-0.93) during 1^st^ lockdown. From 1^st^ re-opening and onwards the level of participation within 180 days returned to pre-pandemic levels (Supplementary Table 5).

The participation in cervical cancer screening within 365 days among women invited at the start of the pandemic was only slightly reduced (Figure 1) reflected in overall PRs of 0.97 (95% CI: 0.96-0.98) during both pre-lockdown and 1^st^ lockdown where after the participation increased to the same level as before the pandemic (Table 2).

**Table 2.** Prevalence ratios and 95% confidence intervals of participation in cervical cancer screening in Denmark within 365 days since invitation from 2015 to 2021*.

|  |  | Pre-pandemic<br>(01Jan2015-31Jan2020) |  | Pre-lockdown<br>(01Feb2020-10Mar2020) |  | 1st lockdown<br>(11Mar2020-15Apr2020) |  | 1st re-opening<br>(16Apr2020-15Dec2020) |  | 2nd lockdown<br>(16Dec2020-31Dec2020) |  |
| --- | --- | --- | --- | --- | --- | --- | --- | --- | --- | --- | --- |
|  |  | N=1641199 |  | N=41876 |  | N=31255 |  | N=223386 |  | N=69729 |  |
|  | N | PR | [95%CI] | PR | [95%CI] | PR | [95%CI] | PR | [95%CI] | PR | [95%CI] |
| **Overall** | 2220000 | 1.00 | - | 0.97 | [0.96; 0.98] | 0.97 | [0.96; 0.98] | 1.01 | [1.00; 1.01] | 0.99 | [0.97; 1.00] |
| **Age at invitation** |  |  |  |  |  |  |  |  |  |  |  |
| 23-29 years | 535235 | 1.00 | - | 0.96 | [0.94; 0.98] | 0.98 | [0.95; 1.00] | 0.99 | [0.98; 1.00] | 1.00 | [0.96; 1.04] |
| 30-39 years | 559296 | 1.00 | - | 0.97 | [0.95; 0.99] | 0.99 | [0.97; 1.01] | 1.01 | [1.00; 1.02] | 1.00 | [0.97; 1.04] |
| 40-49 years | 659365 | 1.00 | - | 0.95 | [0.93; 0.96] | 0.94 | [0.92; 0.95] | 1.00 | [0.99; 1.01] | 0.93 | [0.90; 0.96] |
| 50-59 years | 336786 | 1.00 | - | 1.01 | [0.99; 1.03] | 1.00 | [0.98; 1.02] | 1.02 | [1.01; 1.03] | 1.05 | [1.01; 1.08] |
| 60-64 years | 129318 | 1.00 | - | 0.95 | [0.92; 0.99] | 0.93 | [0.90; 0.97] | 0.98 | [0.96; 1.00] | 0.93 | [0.87; 0.99] |
| **Ethnicity** |  |  |  |  |  |  |  |  |  |  |  |
| Danish descent | 1773076 | 1.00 | - | 0.96 | [0.95; 0.97] | 0.96 | [0.95; 0.97] | 1.00 | [1.00; 1.01] | 0.97 | [0.96; 0.99] |
| Descendant of immigrant | 43902 | 1.00 | - | 0.95 | [0.86; 1.05] | 0.88 | [0.79; 0.98] | 0.97 | [0.92; 1.02] | 0.93 | [0.78; 1.12] |
| Western Immigrant | 120162 | 1.00 | - | 1.03 | [0.98; 1.09] | 1.04 | [0.98; 1.11] | 1.09 | [1.06; 1.12] | 1.06 | [0.95; 1.17] |
| Non-western immigrant | 218679 | 1.00 | - | 0.99 | [0.95; 1.02] | 0.98 | [0.94; 1.02] | 1.04 | [1.02; 1.06] | 1.07 | [1.00; 1.14] |
| **Cohabitation status** |  |  |  |  |  |  |  |  |  |  |  |
| Single | 633585 | 1.00 | - | 0.97 | [0.95; 0.98] | 0.96 | [0.94; 0.98] | 1.02 | [1.01; 1.02] | 0.98 | [0.95; 1.02] |
| Cohabiting | 420793 | 1.00 | - | 0.96 | [0.94; 0.98] | 0.97 | [0.95; 0.99] | 1.00 | [0.99; 1.01] | 1.00 | [0.96; 1.04] |
| Married | 892913 | 1.00 | - | 0.97 | [0.96; 0.98] | 0.97 | [0.96; 0.99] | 1.00 | [1.00; 1.01] | 0.98 | [0.96; 1.01] |
| **Educational level (ISCED)** |  |  |  |  |  |  |  |  |  |  |  |
| ISCED15 level 1-2 | 1297050 | 1.00 | - | 0.96 | [0.95; 0.97] | 0.96 | [0.95; 0.98] | 1.01 | [1.00; 1.01] | 0.99 | [0.97; 1.01] |
| ISCED15 level 3-4 | 529165 | 1.00 | - | 0.96 | [0.95; 0.98] | 0.97 | [0.95; 0.99] | 1.00 | [0.99; 1.01] | 0.97 | [0.94; 1.00] |
| ISCED15 level 5-8 | 319925 | 1.00 | - | 1.00 | [0.98; 1.02] | 0.99 | [0.97; 1.02] | 1.04 | [1.03; 1.05] | 1.06 | [1.01; 1.10] |
| **Disposable income** |  |  |  |  |  |  |  |  |  |  |  |
| Lowest quintile | 419122 | 1.00 | - | 0.95 | [0.93; 0.98] | 0.96 | [0.93; 0.98] | 1.02 | [1.01; 1.04] | 1.02 | [0.97; 1.07] |
| Second quintile | 422225 | 1.00 | - | 0.95 | [0.92; 0.97] | 0.94 | [0.91; 0.96] | 1.00 | [0.99; 1.01] | 1.00 | [0.95; 1.04] |
| Third quintile | 424081 | 1.00 | - | 0.96 | [0.94; 0.98] | 0.95 | [0.93; 0.97] | 0.99 | [0.98; 1.00] | 0.94 | [0.91; 0.98] |
| Fourth quintile | 425069 | 1.00 | - | 0.96 | [0.95; 0.98] | 0.96 | [0.95; 0.98] | 1.00 | [0.99; 1.01] | 0.96 | [0.93; 0.99] |
| Highest quintile | 424457 | 1.00 | - | 0.98 | [0.96; 0.99] | 0.96 | [0.95; 0.98] | 1.00 | [0.99; 1.00] | 0.97 | [0.95; 1.00] |
| **Healthcare usage** |  |  |  |  |  |  |  |  |  |  |  |
| Rare | 439469 | 1.00 | - | 0.97 | [0.95; 0.99] | 0.97 | [0.95; 1.00] | 1.02 | [1.01; 1.03] | 0.99 | [0.94; 1.04] |
| Low | 491164 | 1.00 | - | 0.97 | [0.95; 0.99] | 0.95 | [0.93; 0.97] | 1.00 | [0.99; 1.01] | 0.95 | [0.91; 0.99] |
| Average | 467980 | 1.00 | - | 0.95 | [0.94; 0.97] | 0.95 | [0.93; 0.97] | 1.00 | [0.99; 1.01] | 1.00 | [0.97; 1.04] |
| High | 404486 | 1.00 | - | 0.97 | [0.95; 0.98] | 0.96 | [0.94; 0.98] | 1.01 | [1.00; 1.02] | 0.99 | [0.96; 1.03] |
| Frequent | 416901 | 1.00 | - | 0.97 | [0.95; 0.99] | 0.98 | [0.97; 1.00] | 1.00 | [0.99; 1.01] | 0.96 | [0.93; 1.00] |

\* Adjusted for month, year and age at invitation; PR=prevalence ratio; CI=confidence interval; ISCED=International Standard Classification of Education

### Participation during the COVID-19 pandemic by socio-economic variables

Before the pandemic, the participation in cervical cancer screening within 365 days was lowest among the youngest age group (57%), among immigrants (44-50% in immigrants and 38% in descendants of immigrants), among women living alone (56%), among women with the lowest income level (52%) and among women who rarely use the healthcare system (52%) (Supplementary Figures 2-4 and Supplementary Tables 1-3).

During pre-lockdown and 1^st^ lockdown (the start of the pandemic), the participation in screening within 365 days was reduced among women aged 40-49 years old, 60-64 years old, among descendants of immigrants, among women with a low educational level and a low income (Table 2).

### Time to participation

The median time from invitation to participation was 94 days (IQR=42-200) before the pandemic; however, this increased to 120 days (IQR=72-207) among women invited during pre-lockdown and to 122 days (IQR=53-201) during 1^st^ lockdown. Thereafter, the time to participation resumed to 86 days during the 1^st^ re-opening (Table 1).

## DISCUSSION

### Main findings

In this population-based study, comprising 2,220,000 women invited for cervical cancer screening from 2015-2021 (in 1,466,353 individuals), we found a large decline in participation within 90 days since invitation at the start of the pandemic, a smaller decline in participation within 180 days and only a slight reduction in participation within 365 days. The reduction in participation within 365 days was most pronounced among descendants of immigrants, among women with a low educational level and a low income.

### Comparison with previous studies and explanation of findings

In most countries, population-based screening for cervical cancer was halted at the start of the pandemic. This led to pronounced reductions in the number of women screened for cervical cancer during the early phase of the pandemic (3-5). To our knowledge, no studies have described the long-term participation in cervical cancer screening during the pandemic. We found a marked reduction (42% in pre-lockdown and 24% in 1^st^ lockdown) in the short term (within 90 days) cervical cancer screening participation at the start of the pandemic compared with the previous years. This reduction in participation could be explained either by a change in health behaviour or could perhaps reflect inconsistent messages from the health authorities at the start of the pandemic. The screening programme was open and invitations and reminders were sent out; however, the health authorities asked the population to stay at home at the national televised press conferences and at the same time, the College of General Practitioners recommended general practitioners to postpone routine cervical screening samples during a four-week period in March/ April 2020 (1). The inconsistent health messages could thus have led women to not participate in screening. Congruently, a Danish qualitative study found that inconsistent health communication from the authorities led women to postpone or cancel their screening appointments (21). With the longer follow-up time, we observed a less reduced participation (11% in pre-lockdown and 8% in 1^st^ lockdown within 180 days and only 3% in both pre-lockdown and 1^st^ lockdown within 365 days), which was reflected by the longer time to participation (>120 days versus approx. 89 days) at the early phase of the pandemic. The disruption to the cervical cancer screening programme in Denmark thus appear only to have a temporary effect with most women resuming cervical cancer screening with a longer follow-up period. This is in accordance with findings in a qualitative study showing that women were concerned about visiting healthcare settings during the pandemic but were willing to participate when screening programmes resumed (22). In the Danish cervical cancer screening programme, reminders are sent out to non-participants after 3 and 6 months and this could have prompted women postponing or cancelling their screening appointments at the start of the pandemic to participate at a later time point.

The severity of the pandemic and the pandemic response varied across the world with Denmark managing to keep the number of hospitalisations due to COVID-19 at relatively low level (14). The pandemic response in Denmark included periodic lockdowns, extensive COVID-19 testing free-of-charge to the whole population (15) and a high COVID-19 vaccination coverage. The cervical cancer screening participation may therefore be different in other countries with a different pandemic response and a more severe impact of the pandemic.

Women of lower socio-economic status (7) and immigrant women (8, 9) have earlier been shown to have a lower participation in cervical cancer screening. This was evident from our study also in that immigrants, women living alone and women with a low income level had the lowest participation in cervical cancer screening throughout the study period. A concern is that the pandemic may have affected socially disadvantaged individuals disproportionally. We found an overall 3% reduction in participation within 365 days; however, among descendants of immigrants and among women with a low income a 5% reduction was seen and among women with a low educational level a 4% reduction was found. It is therefore important to ensure that all women – regardless of socioeconomic status – resume participation in cervical cancer screening at the aftermath of the pandemic. To our knowledge, our study is the first to describe cervical cancer screening participation during the pandemic according to socio-economic groups.

A few previous studies have examined the participation in cervical cancer screening during the pandemic according to age groups. One study found that women aged 30-39 years old (6) had the lowest participation in screening during the first six months of the pandemic, whilst another study showed that the oldest age groups (50-59 and 60-69 years old) (23) had the lowest cervical cancer screening participation during the first year of the pandemic. Additionally, a study by Castañon et al. estimated that women aged 40-49 years old would have the greatest burden of excess cervical cancer diagnoses due to a delay in screening because of the pandemic (3). We found that women aged 40-49 years old and 60-64 years old had a lower than usual participation in cervical cancer screening at the start of the pandemic. The pandemic thus appears to affect different age groups differently. Women aged 60-64 years old may have been hesitant to come into contact with the healthcare system at the start of the pandemic possibly explaining the lower participation in this age group. Surprisingly, this effect lasted even when examining participation within 365 days since invitation. A concern is therefore that some women did not resume screening even with the longest follow-up time.

### Strengths and limitations

A major strength of the study is the high quality of data covering the entire population of women invited to participate in the cervical screening programme in Denmark. Danish national registers are known to be reliable and to have high completeness (24), which also confers to the Danish Quality Database for Cervical Screening (17). While the quality of the Danish registers is high, some limitations relate to the data e.g., the study did not include data on comorbidities, which may affect participation in screening during the pandemic as individuals with underlying disease where advised to self-isolate at the height of the pandemic. However, as age is strongly associated with the level of comorbidity, the inclusion of age in the statistical model reduces the theoretical impact of comorbidity on the results.

### Implications of the findings

Our findings show that the overall participation in cervical cancer screening was almost at the same level as the previous years when allowing for the longest follow-up time; however, some groups had a slightly lower participation (descendants of immigrants, women with a low educational level and women with a low income) and it is therefore important to ensure that all women re-enter the cervical cancer screening programme at the aftermath of the pandemic. Our results also show that some age groups (women aged 40-49 years old and 60-64 years old) had a lower participation in screening than usual possibly indicating that the restrictions within a society affects different age groups disproportionally. It is thus important to take this information into account when planning a pandemic response and ensure that all women have access to screening.

Contrasting health messages may have been conveyed by the cervical cancer screening programme being open, the general practitioners recommending a postponement of cervical cancer screening tests and at the same time, the health authorities recommending people to stay at home. Inconsistent health communication from the authorities may therefore have led some women to refrain from participating in screening. The health communication therefore needs to be precise and consistent to ensure that all women are well-informed and know when they can safely participate in cervical cancer screening during a pandemic.

## Conclusion

The cervical cancer screening programme continued throughout the COVID-19 pandemic in Denmark. The participation was reduced at the start of the pandemic; however, most women resumed screening with the longest follow-up time although some groups of women had a slightly lower participation than usual.

## Data Availability

Data availability statement In order to comply with the Danish regulations on data privacy, the datasets generated and analysed during this project are not publicly available as the data are stored and maintained electronically at Statistics Denmark, where it only can be accessed by pre-approved researchers using a secure VPN remote access. Furthermore, no data at a personal level nor data not exclusively necessary for publication are allowed to be extracted from the secure data environment at Statistics Denmark. Access to the data can; however, be granted by the authors of the present study upon a reasonable scientific proposal within the boundaries of the present project and for scientific purposes only.

## Supplementary Tables and figures

**Supplementary Figure 1.**
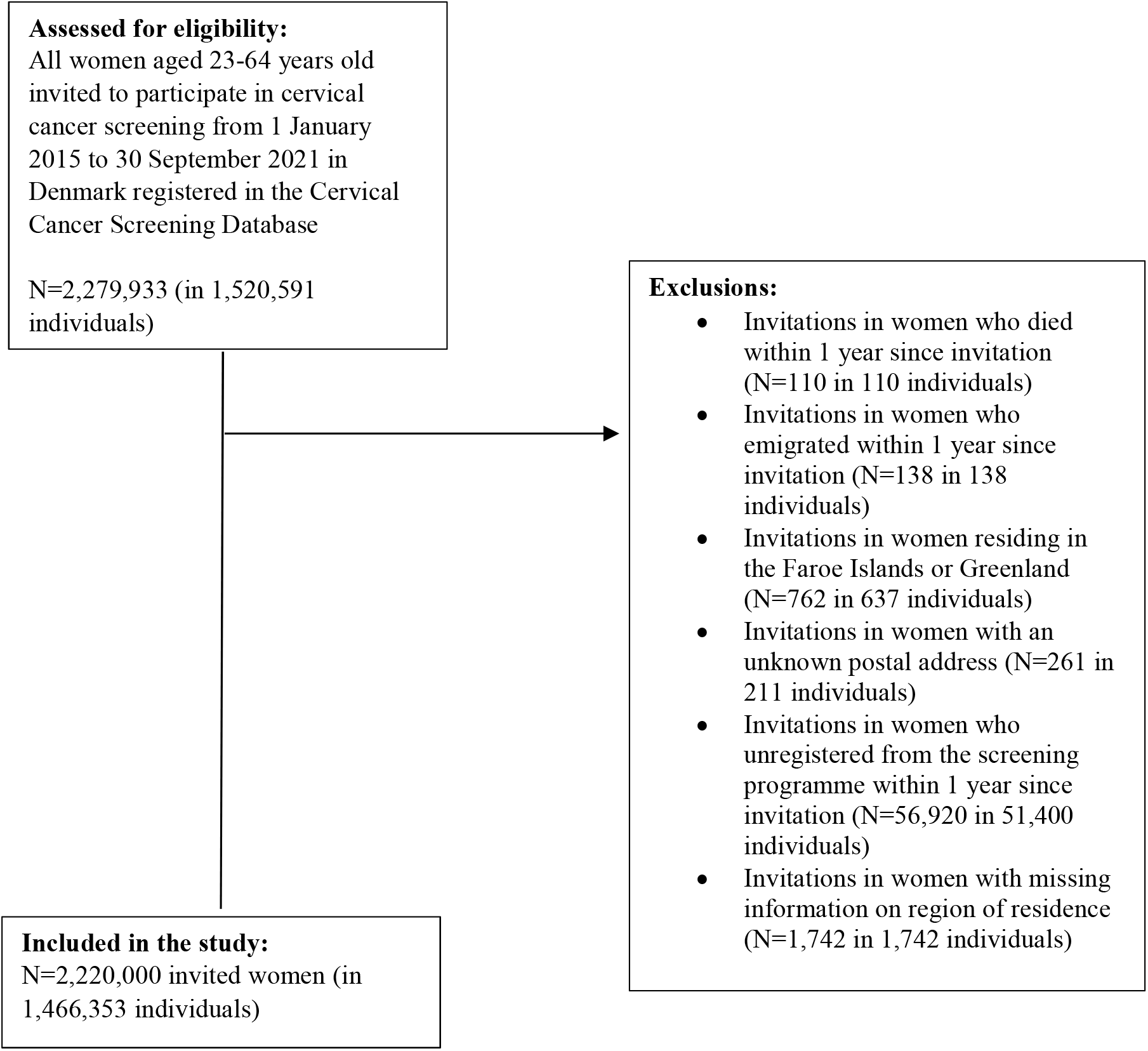
Flow-chart of the study population.

**Supplementary Table 1.** Proportion of women participating in cervical cancer screening in Denmark within 90 days since invitation from 2015 to 2021.

|  | Pre-pandemic<br>(01Jan2015-31Jan2020) | Pre-lockdown<br>(01Feb2020-10Mar2020) | 1st lockdown<br>(11Mar2020-15Apr2020) | 1st re-opening<br>(16Apr2020-15Dec2020) | 2nd lockdown<br>(16Dec2020-27Feb2021) | 2nd re-opening<br>(28Feb2021-30Sep2021) |
| --- | --- | --- | --- | --- | --- | --- |
|  | % | % | % | % | % | % |
| **Total** | 36.2 | 20.6 | 26.7 | 32.0 | 34.3 | 32.3 |
| **Age at invitation** |  |  |  |  |  |  |
| 23-29 years | 28.9 | 16.8 | 21.1 | 25.5 | 27.5 | 24.7 |
| 30-39 years | 33.5 | 19.4 | 25.8 | 29.5 | 30.7 | 29.2 |
| 40-49 years | 39.2 | 21.5 | 28.7 | 35.1 | 37.5 | 36.8 |
| 50-59 years | 41.8 | 25.3 | 30.9 | 38.4 | 41.3 | 39.0 |
| 60-64 years | 47.0 | 25.2 | 34.3 | 42.0 | 45.6 | 43.3 |
| **Ethnicity** |  |  |  |  |  |  |
| Danish descent | 38.9 | 22.4 | 29.0 | 35.3 | 36.8 | 35.7 |
| Descendant of immigrant | 17.6 | 9.4 | 12.4 | 14.8 | 16.4 | 16.5 |
| Western immigrant | 20.9 | 12.2 | 17.0 | 17.4 | 23.7 | 21.6 |
| Non-western immigrant | 26.1 | 13.6 | 16.5 | 21.5 | 23.2 | 22.7 |
| **Cohabitation status** |  |  |  |  |  |  |
| Single | 30.1 | 17.1 | 22.1 | 26.5 | 24.9 | N/A |
| Cohabiting | 34.4 | 20.1 | 26.2 | 31.2 | 31.4 | N/A |
| Married | 41.4 | 23.4 | 30.4 | 36.8 | 35.7 | N/A |
| **Educational level (ISCED)** |  |  |  |  |  |  |
| ISCED15 level 1-2 | 36.4 | 21.0 | 26.2 | 32.3 | 34.4 | 32.9 |
| ISCED15 level 3-4 | 38.0 | 21.1 | 28.4 | 33.8 | 36.7 | 34.5 |
| ISCED15 level 5-8 | 34.3 | 19.0 | 27.3 | 30.4 | 34.6 | 33.2 |
| **Disposable income** |  |  |  |  |  |  |
| Lowest quintile | 26.0 | 14.5 | 17.5 | 23.0 | 24.1 | 22.7 |
| Second quintile | 31.6 | 17.6 | 21.4 | 26.8 | 28.8 | 26.9 |
| Third quintile | 38.1 | 21.6 | 27.6 | 32.5 | 33.1 | 31.8 |
| Fourth quintile | 42.8 | 24.0 | 32.0 | 38.6 | 40.0 | 37.9 |
| Highest quintile | 44.6 | 25.7 | 34.2 | 41.9 | 43.6 | 42.7 |
| **Healthcare usage** |  |  |  |  |  |  |
| Rare | 26.2 | 13.5 | 18.0 | 21.0 | 23.0 | 19.7 |
| Low | 34.2 | 18.4 | 24.2 | 30.6 | 31.8 | 30.8 |
| Average | 37.6 | 21.2 | 27.0 | 34.1 | 35.5 | 34.4 |
| High | 40.3 | 23.9 | 29.9 | 36.4 | 38.7 | 36.8 |
| Frequent | 43.6 | 26.8 | 34.6 | 39.4 | 42.8 | 40.8 |

ISCED=International Standard Classification of Education

**Supplementary Table 2.** Proportion of women participating in cervical cancer screening in Denmark within 180 days since invitation from 2015 to 2021.

|  | Pre-pandemic<br>(01Jan2015-31Jan2020) | Pre-lockdown<br>(01Feb2020-10Mar2020) | 1st lockdown<br>(11Mar2020-15Apr2020) | 1st re-opening<br>(16Apr2020-15Dec2020) | 2nd lockdown<br>(16Dec2020-27Feb2021) | 2nd re-opening<br>(28Feb2021-30Jun2021) |
| --- | --- | --- | --- | --- | --- | --- |
|  | % | % | % | % | % | % |
| **Total** | 53.8 | 44.3 | 46.1 | 49.0 | 50.1 | 49.5 |
| **Age at invitation** |  |  |  |  |  |  |
| 23-29 years | 44.8 | 36.0 | 38.2 | 40.7 | 41.1 | 39.8 |
| 30-39 years | 51.4 | 41.3 | 44.3 | 46.5 | 47.0 | 46.7 |
| 40-49 years | 58.7 | 47.9 | 49.4 | 54.2 | 54.7 | 55.3 |
| 50-59 years | 58.4 | 52.1 | 52.4 | 55.1 | 57.3 | 56.4 |
| 60-64 years | 62.8 | 55.3 | 55.0 | 58.3 | 61.1 | 60.2 |
| **Ethnicity** |  |  |  |  |  |  |
| Danish descent | 57.4 | 47.9 | 49.6 | 53.4 | 53.4 | 53.6 |
| Descendant of immigrant | 28.3 | 21.2 | 20.1 | 25.1 | 24.5 | 26.5 |
| Western immigrant | 33.4 | 27.3 | 30.2 | 30.1 | 35.5 | 35.1 |
| Non-western immigrant | 40.2 | 30.8 | 32.1 | 34.8 | 36.8 | 36.5 |
| **Cohabitation status** |  |  |  |  |  |  |
| Single | 45.3 | 37.4 | 38.6 | 41.4 | 39.3 | N/A |
| Cohabiting | 52.0 | 42.4 | 45.6 | 48.4 | 47.3 | N/A |
| Married | 60.6 | 50.2 | 52.0 | 55.4 | 54.7 | N/A |
| **Educational level (ISCED)** |  |  |  |  |  |  |
| ISCED15 level 1-2 | 53.8 | 44.5 | 45.5 | 49.3 | 50.0 | 49.8 |
| ISCED15 level 3-4 | 56.0 | 46.0 | 47.8 | 51.2 | 52.9 | 52.1 |
| ISCED15 level 5-8 | 52.1 | 41.9 | 47.2 | 47.3 | 51.8 | 51.4 |
| **Disposable income** |  |  |  |  |  |  |
| Lowest quintile | 40.6 | 31.7 | 32.9 | 37.2 | 36.7 | 36.3 |
| Second quintile | 46.8 | 37.3 | 37.6 | 41.7 | 42.8 | 41.1 |
| Third quintile | 55.5 | 45.4 | 46.4 | 49.5 | 48.8 | 48.2 |
| Fourth quintile | 62.8 | 51.9 | 53.7 | 58.1 | 57.0 | 56.3 |
| Highest quintile | 65.9 | 55.8 | 58.5 | 62.4 | 62.3 | 63.1 |
| **Healthcare usage** |  |  |  |  |  |  |
| Rare | 41.2 | 31.7 | 34.5 | 34.7 | 35.2 | 33.5 |
| Low | 52.0 | 42.7 | 43.3 | 48.1 | 47.9 | 47.9 |
| Average | 56.1 | 45.8 | 47.4 | 52.0 | 52.5 | 52.0 |
| High | 58.9 | 49.4 | 50.7 | 55.0 | 55.9 | 55.0 |
| Frequent | 61.5 | 52.8 | 55.0 | 57.3 | 59.0 | 59.3 |

ISCED=International Standard Classification of Education

**Supplementary Table 3.** Proportion of women participating in cervical cancer screening in Denmark within 365 days since invitation from 2015 to 2021.

|  | Pre-pandemic<br>(01Jan2015-31Jan2020) | Pre-lockdown<br>(01Feb2020-10Mar2020) | 1st lockdown<br>(11Mar2020-15Apr2020) | 1st re-opening<br>(16Apr2020-15Dec2020) | 2nd lockdown<br>(16Dec2020-31Dec2020) |
| --- | --- | --- | --- | --- | --- |
|  | % | % | % | % | % |
| **Total** | 64.9 | 60.5 | 60.2 | 60.3 | 57.3 |
| **Age at invitation** |  |  |  |  |  |
| 23-29 years | 57.0 | 51.5 | 52.5 | 52.5 | 50.3 |
| 30-39 years | 64.3 | 59.0 | 59.9 | 59.7 | 57.3 |
| 40-49 years | 69.9 | 64.8 | 64.0 | 65.5 | 59.2 |
| 50-59 years | 66.4 | 66.4 | 64.2 | 63.6 | 63.2 |
| 60-64 years | 69.0 | 68.1 | 64.7 | 65.9 | 59.4 |
| **Ethnicity** |  |  |  |  |  |
| Danish descent | 68.7 | 64.7 | 64.1 | 65.1 | 61.8 |
| Descendant of immigrant | 37.8 | 33.7 | 30.5 | 34.1 | 28.9 |
| Western immigrant | 43.8 | 41.7 | 42.6 | 41.3 | 38.1 |
| Non-western immigrant | 50.5 | 44.9 | 44.3 | 44.6 | 42.6 |
| **Cohabitation status** |  |  |  |  |  |
| Single | 56.0 | 52.2 | 51.5 | 52.1 | 47.9 |
| Cohabiting | 64.4 | 59.9 | 60.9 | 61.1 | 59.1 |
| Married | 71.4 | 66.9 | 66.4 | 66.7 | 64.1 |
| **Educational level (ISCED)** |  |  |  |  |  |
| ISCED15 level 1-2 | 64.8 | 60.4 | 59.6 | 60.6 | 56.8 |
| ISCED15 level 3-4 | 67.0 | 62.6 | 62.0 | 62.5 | 61.0 |
| ISCED15 level 5-8 | 63.8 | 58.9 | 61.2 | 59.4 | 59.4 |
| **Disposable income** |  |  |  |  |  |
| Lowest quintile | 52.0 | 46.8 | 46.5 | 48.7 | 46.1 |
| Second quintile | 57.5 | 51.9 | 50.7 | 52.6 | 50.6 |
| Third quintile | 66.4 | 61.1 | 60.2 | 60.5 | 55.7 |
| Fourth quintile | 73.9 | 68.8 | 68.7 | 69.5 | 65.1 |
| Highest quintile | 77.4 | 74.5 | 73.4 | 74.3 | 71.5 |
| **Healthcare usage** |  |  |  |  |  |
| Rare | 51.5 | 46.7 | 46.4 | 45.3 | 42.2 |
| Low | 63.6 | 59.9 | 58.4 | 59.7 | 54.9 |
| Average | 67.6 | 62.6 | 62.1 | 63.7 | 62.4 |
| High | 70.2 | 66.0 | 65.3 | 66.8 | 64.1 |
| Frequent | 72.2 | 68.1 | 68.9 | 68.5 | 64.4 |

ISCED=International Standard Classification of Education

**Supplementary Table 4.**
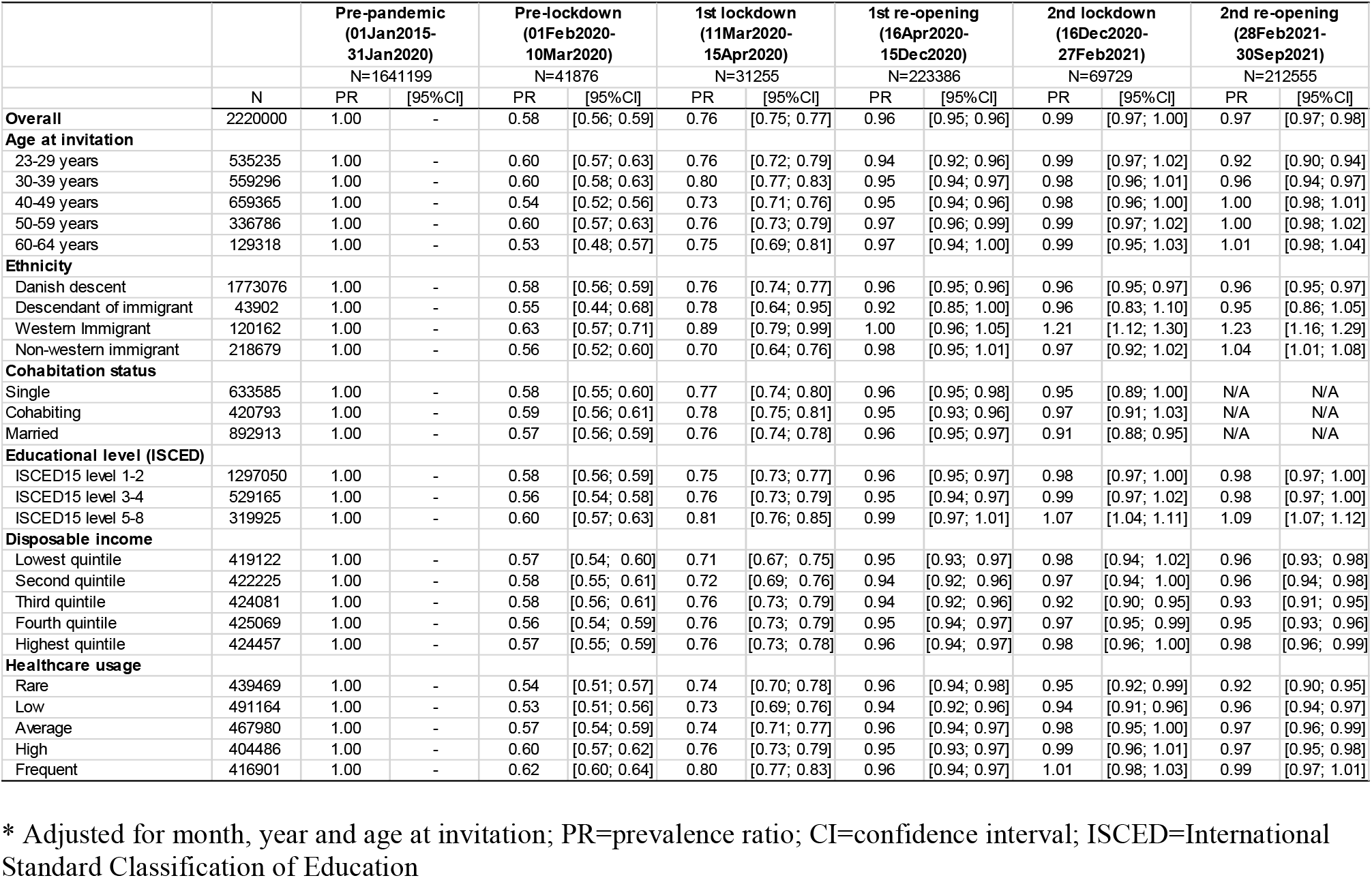
Prevalence ratios and 95% confidence intervals of participation in cervical cancer screening in Denmark within 90 days since invitation from 2015 to 2021.

**Supplementary Table 5.** Prevalence ratios and 95% confidence intervals of participation in cervical cancer screening in Denmark within 180 days since invitation from 2015 to 2021*.

|  |  | Pre-pandemic<br>(01Jan2015-31Jan2020) |  | Pre-lockdown<br>(01Feb2020-10Mar2020) |  | 1st lock-down<br>(11Mar2020-15Apr2020) |  | 1st re-opening<br>(16Apr2020-15Dec2020) |  | 2nd lock-down<br>(16Dec2020-27Feb2021) |  | 2nd re-opening<br>(28Feb2021-30June2021) |  |
| --- | --- | --- | --- | --- | --- | --- | --- | --- | --- | --- | --- | --- | --- |
|  |  | N=1641199 |  | N=41876 |  | N=31255 |  | N=223386 |  | N=69729 |  | N=212555 |  |
|  | N | PR | [95%CI] | PR | [95%CI] | PR | [95%CI] | PR | [95%CI] | PR | [95%CI] | PR | [95%CI] |
| **Overall** | 2220000 | 1.00 | - | 0.89 | [0.88; 0.90] | 0.92 | [0.91; 0.93] | 0.99 | [0.98; 0.99] | 1.02 | [1.01; 1.03] | 1.00 | [1.00; 1.01] |
| **Age at invitation** |  |  |  |  |  |  |  |  |  |  |  |  |  |
| 23-29 years | 535235 | 1.00 | - | 0.89 | [0.86; 0.91] | 0.92 | [0.89; 0.95] | 0.98 | [0.96; 0.99] | 1.02 | [1.00; 1.04] | 0.98 | [0.96; 0.99] |
| 30-39 years | 559296 | 1.00 | - | 0.89 | [0.86; 0.91] | 0.94 | [0.91; 0.96] | 0.99 | [0.98; 1.00] | 1.03 | [1.01; 1.05] | 1.01 | [0.99; 1.02] |
| 40-49 years | 659365 | 1.00 | - | 0.86 | [0.84; 0.88] | 0.88 | [0.86; 0.90] | 0.98 | [0.97; 0.99] | 1.00 | [0.99; 1.02] | 0.99 | [0.98; 1.01] |
| 50-59 years | 336786 | 1.00 | - | 0.92 | [0.90; 0.95] | 0.95 | [0.92; 0.97] | 1.00 | [0.99; 1.02] | 1.03 | [1.01; 1.05] | 1.03 | [1.01; 1.05] |
| 60-64 years | 129318 | 1.00 | - | 0.88 | [0.84; 0.92] | 0.91 | [0.86; 0.95] | 0.97 | [0.95; 0.99] | 1.00 | [0.97; 1.04] | 0.99 | [0.97; 1.02] |
| **Ethnicity** |  |  |  |  |  |  |  |  |  |  |  |  |  |
| Danish descent | 1773076 | 1.00 | - | 0.88 | [0.87; 0.89] | 0.91 | [0.90; 0.92] | 0.99 | [0.98; 0.99] | 0.99 | [0.98; 1.00] | 0.98 | [0.98; 0.99] |
| Descendant of immigrant | 43902 | 1.00 | - | 0.84 | [0.73; 0.96] | 0.82 | [0.70; 0.95] | 0.96 | [0.91; 1.02] | 0.93 | [0.84; 1.04] | 0.96 | [0.88; 1.05] |
| Western Immigrant | 120162 | 1.00 | - | 0.92 | [0.86; 0.99] | 1.01 | [0.93; 1.09] | 1.06 | [1.03; 1.10] | 1.18 | [1.11; 1.24] | 1.18 | [1.12; 1.24] |
| Non-western immigrant | 218679 | 1.00 | - | 0.89 | [0.84; 0.93] | 0.92 | [0.88; 0.98] | 1.02 | [1.00; 1.04] | 1.08 | [1.04; 1.12] | 1.09 | [1.05; 1.12] |
| **Cohabitation status** |  |  |  |  |  |  |  |  |  |  |  |  |  |
| Single | 633585 | 1.00 | - | 0.88 | [0.86; 0.90] | 0.91 | [0.88; 0.93] | 1.00 | [0.99; 1.01] | 1.03 | [0.98; 1.07] | N/A | N/A |
| Cohabiting | 420793 | 1.00 | - | 0.87 | [0.85; 0.90] | 0.92 | [0.89; 0.95] | 0.98 | [0.97; 0.99] | 1.01 | [0.97; 1.06] | N/A | N/A |
| Married | 892913 | 1.00 | - | 0.89 | [0.88; 0.90] | 0.92 | [0.90; 0.94] | 0.99 | [0.98; 0.99] | 1.01 | [0.98; 1.03] | N/A | N/A |
| **Educational level (ISCED)** |  |  |  |  |  |  |  |  |  |  |  |  |  |
| ISCED15 level 1-2 | 1297050 | 1.00 | - | 0.89 | [0.87; 0.90] | 0.91 | [0.89; 0.92] | 0.99 | [0.98; 1.00] | 1.02 | [1.00; 1.03] | 1.00 | [0.99; 1.01] |
| ISCED15 level 3-4 | 529165 | 1.00 | - | 0.88 | [0.86; 0.90] | 0.90 | [0.88; 0.93] | 0.98 | [0.97; 0.99] | 1.02 | [1.00; 1.04] | 1.02 | [1.01; 1.03] |
| ISCED15 level 5-8 | 319925 | 1.00 | - | 0.89 | [0.86; 0.92] | 0.97 | [0.94; 1.00] | 1.02 | [1.01; 1.04] | 1.11 | [1.09; 1.14] | 1.08 | [1.06; 1.10] |
| **Disposable income** |  |  |  |  |  |  |  |  |  |  |  |  |  |
| Lowest quintile | 419122 | 1.00 | - | 0.86 | [0.83; 0.89] | 0.89 | [0.85; 0.92] | 1.00 | [0.98; 1.01] | 1.02 | [1.00; 1.05] | 1.00 | [0.98; 1.02] |
| Second quintile | 422225 | 1.00 | - | 0.87 | [0.84; 0.90] | 0.88 | [0.85; 0.91] | 0.98 | [0.96; 0.99] | 1.01 | [0.99; 1.04] | 0.98 | [0.96; 1.00] |
| Third quintile | 424081 | 1.00 | - | 0.89 | [0.86; 0.91] | 0.90 | [0.88; 0.93] | 0.97 | [0.96; 0.98] | 0.98 | [0.96; 1.00] | 0.96 | [0.95; 0.98] |
| Fourth quintile | 425069 | 1.00 | - | 0.88 | [0.86; 0.90] | 0.90 | [0.88; 0.93] | 0.98 | [0.97; 0.99] | 0.99 | [0.97; 1.01] | 0.97 | [0.95; 0.98] |
| Highest quintile | 424457 | 1.00 | - | 0.88 | [0.87; 0.90] | 0.92 | [0.90; 0.94] | 0.98 | [0.97; 0.99] | 0.99 | [0.98; 1.01] | 0.99 | [0.98; 1.00] |
| **Healthcare usage** |  |  |  |  |  |  |  |  |  |  |  |  |  |
| Rare | 439469 | 1.00 | - | 0.87 | [0.84; 0.90] | 0.94 | [0.91; 0.98] | 1.01 | [0.99; 1.02] | 0.99 | [0.97; 1.02] | 0.97 | [0.95; 0.99] |
| Low | 491164 | 1.00 | - | 0.87 | [0.85; 0.89] | 0.88 | [0.86; 0.91] | 0.98 | [0.96; 0.99] | 0.99 | [0.97; 1.01] | 0.98 | [0.96; 1.00] |
| Average | 467980 | 1.00 | - | 0.87 | [0.85; 0.89] | 0.90 | [0.87; 0.92] | 0.98 | [0.97; 0.99] | 1.01 | [0.99; 1.03] | 0.99 | [0.98; 1.01] |
| High | 404486 | 1.00 | - | 0.89 | [0.87; 0.91] | 0.91 | [0.88; 0.93] | 0.99 | [0.98; 1.00] | 1.02 | [1.00; 1.04] | 0.99 | [0.98; 1.01] |
| Frequent | 416901 | 1.00 | - | 0.91 | [0.89; 0.93] | 0.93 | [0.91; 0.96] | 0.98 | [0.97; 0.99] | 1.02 | [1.01; 1.04] | 1.02 | [1.01; 1.03] |

\* Adjusted for month, year and age at invitation; PR=prevalence ratio; CI=confidence interval; ISCED=International Standard Classification of Education

**Supplementary Figure 2.**
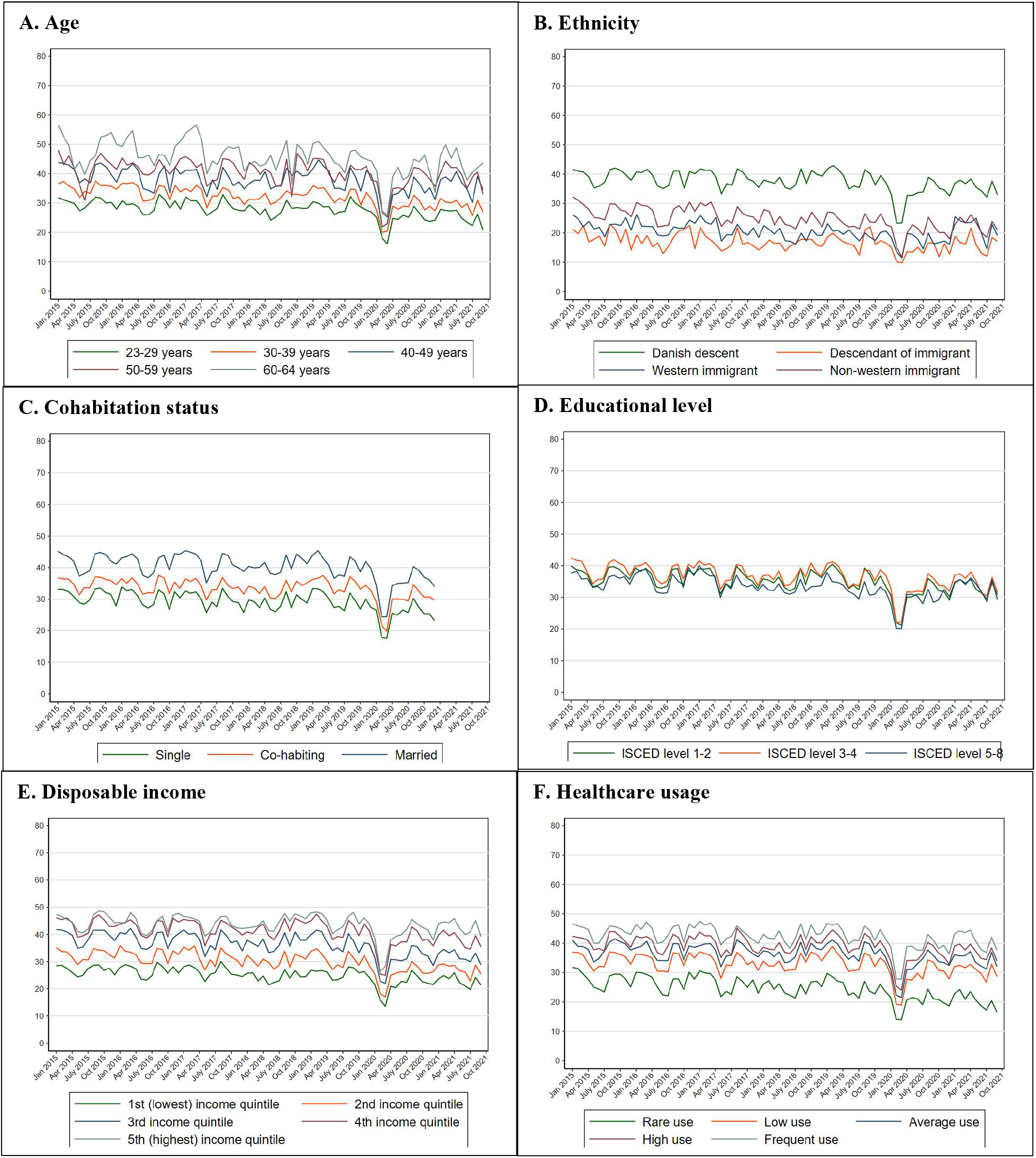
Proportion of women participating in cervical cancer screening in Denmark within 90 days since invitation from 2015 to 2021 stratified by the explanatory variables.

**Supplementary Figure 3.**
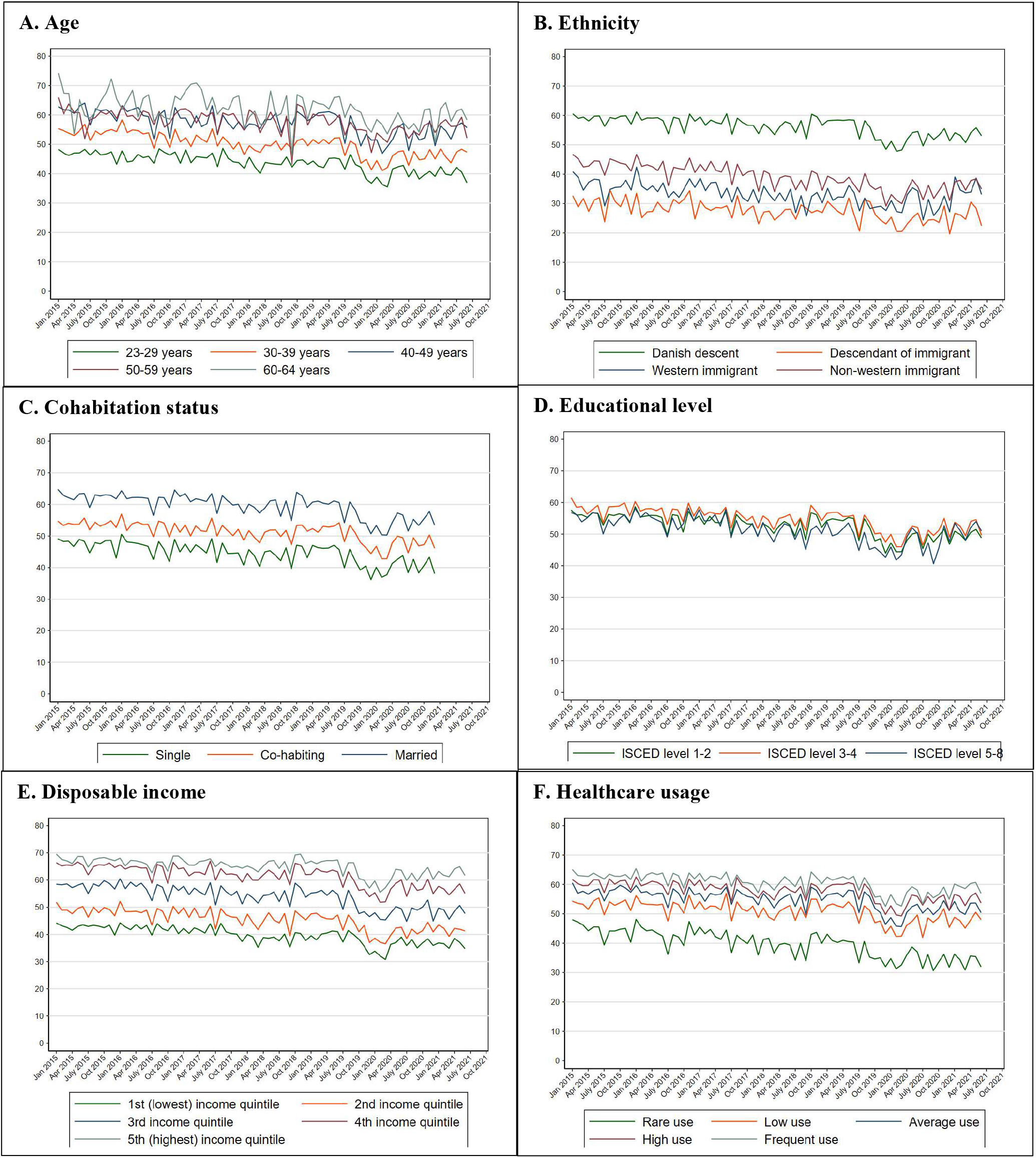
Proportion of women participating in cervical cancer screening in Denmark within 180 days since invitation from 2015 to 2021 stratified by the explanatory variables.

**Supplementary Figure 4.**
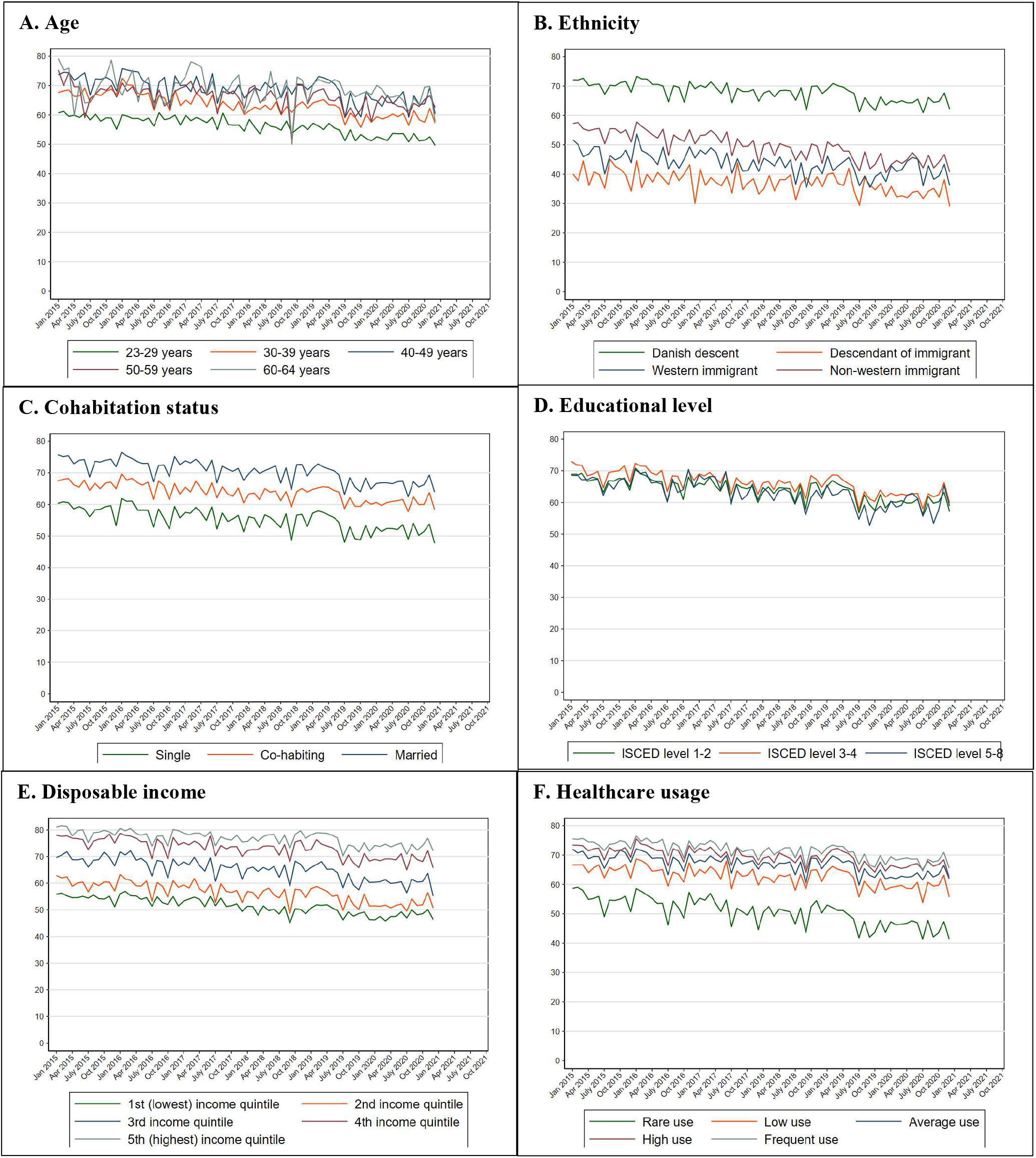
Proportion of women participating in cervical cancer screening in Denmark within 365 days since invitation from 2015 to 2021 stratified by the explanatory variables.

## Authorship

1. TBO, HJ, HM, JWJ, MW and BA designed the study. TBO and HJ acquired the data; HJ analysed the data. All authors contributed to the interpretation of the data.
2. TBO drafted the article. All authors revised the article critically for important intellectual content.
3. All authors approved the final version of the article to be published.
4. All authors agree to be accountable for all aspects of the work in ensuring that questions related to the accuracy or integrity of any part of the work are appropriately investigated and resolved.

## Acknowledgements

We would like to thank Flemming Bro, MD, PhD, GP, Senior researcher from the Research Unit for General Practice, Aarhus for his valuable comments to the manuscript.

## Materials availability statement

Not applicable.

## Data availability statement

In order to comply with the Danish regulations on data privacy, the datasets generated and analysed during this project are not publicly available as the data are stored and maintained electronically at Statistics Denmark, where it only can be accessed by pre-approved researchers using a secure VPN remote access. Furthermore, no data at a personal level nor data not exclusively necessary for publication are allowed to be extracted from the secure data environment at Statistics Denmark. Access to the data can; however, be granted by the authors of the present study upon a reasonable scientific proposal within the boundaries of the present project and for scientific purposes only.

